# The ratio of monocyte to apolipoprotein A1 is an independent predictor of hepatocellular carcinoma: a retrospective study

**DOI:** 10.1101/2025.05.29.25328608

**Authors:** Cheng Su, Xuefeng Zhu, Zhongyuan Lin, Fu Wei, Ling Li

## Abstract

**Objective:** The ratio of monocyte to apolipoprotein A1 (MAR) is a new diagnostic indicator of some chronic diseases, but there have been no reports on hepatocellular carcinoma (HCC). This study aimed to explore the predictive value of MAR in HCC.

**Methods:** A total of 270 patients with HCC, 540 hepatitis B patients and 540 healthy volunteers were included. Laboratory data including monocyte counts and apolipoprotein A1 levels were retrospectively collected from electronic medical records. The truncation value, area under the curve (AUC) and Youden index of each significant variable were calculated using the receiver operating characteristic curve (ROC). The clinical value of MAR was analyzed by univariate and multivariate logistic regression. Correlation heatmap was used to analyze the correlation between each index and MAR.

**Results:** MAR level was the highest in the HCC, followed by hepatitis B disease, and the lowest in the healthy volunteer (*P* < 0.001), and it was found to be an independent risk factor for HCC, also an indicator to distinguish HCC from hepatitis B disease. The cut-off value of MAR for distinguishing HCC from hepatitis B was 0.53; the AUC was 0.762, while the cut-off value in the HCC and the healthy volunteer groups was 0.62; the AUC was 0.829. The increase in MAR level showed that the risk of HCC was 6.001 times higher than that of healthy volunteer (*P* < 0.001). MAR was correlated with indicators, such as lymphocytes (r = -0.075, *P* < 0.05), carcinoembryonic antigen (r = 0.171, *P* < 0.001), albumin (r = -0.445, *P* < 0.001).

**Conclusions:** MAR is a new indicator for the differential diagnosis of HCC and hepatitis B disease, and also an independent risk factor for HCC. This is the first study to explore the clinical value of MAR in HCC.

## Introduction

Liver cancer is one of the most common malignant tumors of the digestive tract worldwide, with a 5-year survival of 18%[1]. Hepatocellular Carcinoma (HCC) accounts for 90% of the cases[2]. According to the latest statistics, approximately 50–80% of HCC cases worldwide are caused by chronic hepatitis B virus (HBV) infection[3]. HBV infection can promote hepatocellular carcinogenesis through direct or indirect mechanisms. The liver microenvironment changes through HBV infection-induced chronic inflammation, the interaction between the virus and innate and adaptive immune cells, which helps the virus evade immune surveillance and promote disease evolution from inflammation to tumor formation. The early stage of HCC is insidious, and most patients are already in the late stage when symptoms appear. Therefore, early screening and diagnosis of HCC are of great clinical significance for the development of suitable treatment programs, and determine more reliable hematological predictors that are closely related to the characteristics of HCC. About 15% of the global cancer burden is attributable to infectious agents, and inflammation is a major component of these chronic infections. The relationship between cancer-related chronic inflammation, dyslipidemia, and malignant tumors has attracted much attention nowadays[4,5]. Additionally, chronic inflammation is closely related to the occurrence and development of tumors[6]. Inflammation provides a good environment for oncogenesis. Tumour-associated macrophages (TAM) are a major component of the infiltrate of most, if not all, tumours[7]. TAM are a subpopulation of monocytes that originates in the circulating blood and is activated around the tumor due to the release of tumor chemokines[8]. In recent years, inflammatory markers such as monocyte count have been found to be associated with HCC risk. Monocytes (M) are the innate immune cells of the mononuclear phagocyte system in chronic inflammation, and have become an important regulator of cancer development and progression[9].

The presence of lipids in the human body mainly comes from the diet and the synthesis of liver cells. The change of the local microenvironment of tumor may cause lipid metabolism disorder[10]. ApoA1 is the main apolipoprotein in HDL and the largest component of ApoA family, which plays a key role in blood lipids[11]. It has been shown that dyslipidemia can increase the risk of tumor and is closely related to the prognosis and development of diseases, such as prostate cancer, colorectal cancer and HCC[12-14]. MAR, obtained from the ratio of M to ApoA1, is a new diagnostic indicator of some chronic diseases in recent years, such as type-2 diabetes and breast cancer[15,16], but there have been no reports on HCC.

As the liver is the main organ for metabolism, we are curious whether MAR indicators are meaningful in the diagnosis of HCC. Therefore, the purpose of this study is to explore the clinical diagnostic value of MAR in HCC. We retrospectively collected the hematological test results of 270 HCC patients, 540 hepatitis B patients and 540 healthy controls, using statistical analysis software to analyze the clinical diagnostic value of MAR in HCC.

## Patients and Methods

### Patients

HCC and HBV patients hospitalized in the Department of Gastroenterology of the People’s Hospital of Guangxi Zhuang Autonomous Region from January 1, 2020 to November 30, 2024 were selected as the research objects. The baseline data, laboratory test indicators and clinical history were collected retrospectively. The inclusion criteria of patients: 1) Complete clinical data; 2) HCC patients who meet the diagnostic criteria of the 2018 Chinese Society of Clinical Oncology (CSCO) Guidelines for the Diagnosis and Treatment of HCC; 3) No anti-cancer treatment. 4) Patients with hepatitis B diagnosed in accordance with the 2019 Guidelines for the Prevention and Treatment of Chronic Hepatitis B. Exclusion criteria: 1) Patients with liver metastases or secondary tumors; 2) Simultaneously suffering from other tumors;3) Patients with a history of major organ transplantation or other serious illnesses; 4) Merge hepatitis A, C, E virus or other viral infections such as HIV; 5) Those with unclear diagnosis. The healthy people without liver cancer and hepatitis B who were examined in the physical examination center of our hospital at the same time were selected as the control group. This study has been approved by the Ethics Committee of our hospital, meets the ethical requirements specified by the Ethics Committee (Ethical code: KY-IIT-2025-46), and has obtained written informed consent from the subjects (or guardians).

## Methods

### Laboratory indicators

Fasting venous blood of patients in the morning was collected and placed into EDTA-K2 anticoagulant tube (2ml) and procoagulant tube (5ml). These laboratory parameters, such as lymphocytes (L), platelet (PLT), monocyte (M), ApoA1, were tested by Sysmex (XN9000 automatic blood analyzer). Serum carcinoembryonic antigen (CEA) level was measured by Roche cobas 801 automatic chemiluminescence immunoassay analyzer. Apolipoprotein B (ApoB), albumin (ALB) and alkaline phosphatase (ALP) were determined by Beckman coulter AU5800 automatic biochemical analyzer. Alpha-fetoprotein (AFP) was measured by Beckman coulter UniCel Dxl 800 Access immune analyzer. These instruments were used after daily quality control inspections in strict accordance with operation. PLR is the ratio of platelet to lymphocyte; LAR is the ratio of lymphocyte to ApoA1; LMR is the ratio of lymphocyte to monocyte; PAR is the ratio of platelet to ApoA1; BAR is the ratio of ApoB to ApoA1; MAR is the ratio of monocyte to ApoA1.

### Statistical Analysis

In this study, the HCC group was matched with hepatitis B patients and healthy people by 1:2 propensity score matching (PSM) method using R software. There was no significant difference in age. SPSS 26.0 software (IBM corp, Armonk, NY) was used for data analysis. Measurement data with normal distribution were represented as mean ± SD, non-normal distribution were expressed as median (P_25_, P_75_), and Kruskal-Wallis H-test was used for comparison between groups. The truncation value, area under the curve (AUC) and Youden index of each significant variable were calculated using the receiver operating characteristic curve (ROC). Univariate and multivariate logistic regression was used to analyze the risk factors of HCC. Correlation heatmap was used to analyze the correlation between each index and the MAR, and the correlation coefficient r value was calculated. A two-tailed *P* value of no more than 0.05 was considered statistically significant. Furthermore, ROC curve were performed using GraphPad Prism 8 (GraphPad Software, San Diego, CA, USA) software.

## Results

The information about the subjects available was retrospectively collected from the medical electronic recording system. A total of 270 HCC patients, 540 hepatitis B patients and 540 healthy controls were enrolled in this study. Their age is mainly concentrated around 57 years old.

### Difference Analysis of Three Groups

The comparison of hematological indicators among the HCC group, the hepatitis B group and the healthy volunteer group showed that there were statistically significant differences in PLT (*P* < 0.001), L (*P* < 0.001), M (*P* < 0.001), CEA (*P* < 0.001), ALB (*P* < 0.001), ApoA1 (*P* < 0.001), ApoB (*P* < 0.001), ALP (*P* < 0.001), PLR (*P* < 0.001), PAR (*P* < 0.001), BAR (*P* < 0.001), LAR (*P* < 0.001), LMR (*P* < 0.001), MAR (*P* < 0.001) among the three groups (Table 1). Subsequently, the comparison between the two groups showed that PLT, L, M, ALB, ApoA1, ApoB, PLR, PAR, BAR, LAR, LMR was notably different between healthy volunteers and hepatitis B groups (*P* < 0.05). There were significant differences in PLT, L, M, CEA, ALB, ApoA1, ApoB, ALP, BAR, LAR, LMR, MAR between healthy volunteers and HCC groups (*P* < 0.05). Moreover, there were significant difference in L, M, CEA, ALB, ApoA1, ALP, PLR, PAR, BAR, LMR, MAR between hepatitis B group and HCC group (*P* < 0.05). Among them, MAR level was the highest in the HCC group (0.92), followed by hepatitis B group (0.38), and the lowest in the healthy volunteer group (0.36), as shown in Table 1.

**Table 1.** Comparison of Hematological Parameters Among Patients with Hepatocellular Carcinoma, Those with hepatitis B patients and Healthy Volunteers.

| Group | Healthy Volunteers | hepatitis B patients | Hepatocellular Carcinoma | H | *P* |
| --- | --- | --- | --- | --- | --- |
| N | 540 | 540 | 270 |  |  |
| Age (years) | 57(50, 66) | 58(50, 65) | 57(50, 66) | 0.233 | 0.890 |
| PLT ($\times 10^9/L$) | 263(224, 305) | 172(101, 224)<sup>a</sup> | 151(98, 226)<sup>b</sup> | 406.897 | 0.000 |
| L ($\times 10^9/L$) | 2.20(1.76, 2.62) | 1.52(1.05, 1.94)<sup>a</sup> | 1.06(0.76, 1.51)<sup>bc</sup> | 413.560 | 0.000 |
| M ($\times 10^9/L$) | 0.50(0.40, 0.60) | 0.42(0.32, 0.57)<sup>a</sup> | 0.63(0.43, 0.95)<sup>bc</sup> | 121.623 | 0.000 |
| CEA ($\mu g/L$) | 1.88(1.41, 2.57) | 2.04(1.20, 3.69) | 2.36(1.52, 3.99)<sup>bc</sup> | 22.494 | 0.000 |
| AFP ($\mu g/L$) | 2.96(2.28, 3.95) | 2.91(1.81, 6.33) | 7.25(0.96, 95.91) | 3.493 | 0.174 |
| ALB(g/L) | 45.00(43.10, 47.42) | 37.10(30.60, 40.30)<sup>a</sup> | 31.00(27.00, 35.45)<sup>bc</sup> | 772.356 | 0.000 |
| ApoA1 (g/L) | 1.31(1.20, 1.46) | 1.10(0.88, 1.32)<sup>a</sup> | 0.75(0.53, 1.03)<sup>bc</sup> | 420.352 | 0.000 |
| ApoB (g/L) | 0.98(0.85, 1.12) | 0.85(0.67, 1.05)<sup>a</sup> | 0.83(0.63, 1.05)<sup>b</sup> | 69.593 | 0.000 |
| ALP (U/L) | 75(63, 89) | 75(60, 104) | 126(96, 213)<sup>bc</sup> | 246.483 | 0.000 |
| PLR | 121.05(97.82, 151.85) | 108.37(76.06, 142.97)<sup>a</sup> | 137.98(89.66, 216.80)<sup>c</sup> | 48.371 | 0.000 |
| PAR | 196.97(166.60, 240.89) | 152.18(103.88, 207.09)<sup>a</sup> | 205.54(113.11, 356.39)<sup>c</sup> | 99.343 | 0.000 |
| BAR | 0.74(0.61, 0.86) | 0.79(0.61, 1.00)<sup>a</sup> | 1.07(0.77, 1.62)<sup>bc</sup> | 137.446 | 0.000 |
| LAR | 1.65(1.30, 2.03) | 1.38(0.97, 1.88)<sup>a</sup> | 1.52(0.93, 2.19)<sup>b</sup> | 35.553 | 0.000 |
| LMR | 4.40(3.48, 5.60) | 3.37(2.26, 4.86)<sup>a</sup> | 1.61(1.06, 2.58)<sup>bc</sup> | 386.867 | 0.000 |
| MAR | 0.36(0.29, 0.47) | 0.38(0.26, 0.60)<sup>a</sup> | 0.92(0.47, 1.60)<sup>bc</sup> | 232.113 | 0.000 |

Notes: <sup>a</sup> Healthy Volunteers vs hepatitis B patients, $P < 0.05$ ; <sup>b</sup> Healthy Volunteers vs Hepatocellular Carcinoma, $P < 0.05$ ; <sup>c</sup> hepatitis B patients vs Hepatocellular Carcinoma, $P < 0.05$ . *P* value is analyzed by Kruskal-Wallis H-test.
Abbreviations: AFP, Alpha-fetoprotein; ALB, albumin; ApoA1, apolipoprotein A1; ApoB, Apolipoprotein B; ALP, alkaline phosphatase; BAR, ApoB to ApoA1; CEA, carcinoembryonic antigen; L, lymphocytes; LAR, lymphocyte to ApoA1; LMR, lymphocyte to monocyte; M, Monocytes; MAR, monocyte to ApoA1; PLT, platelet; PLR, platelet to lymphocyte; PAR, platelet to ApoA1.

### Calculation of Cut-off Value of MAR

Draw ROC curves and calculate the cut-off values and AUC for NLR, PLR, NAR, PAR, LAR, LMR, and MAR (Table 2). The cut-off value of MAR for distinguishing HCC from hepatitis B was 0.53; the area under the curve was 0.762, while the cut-off value in the HCC and the healthy volunteer groups was 0.62; the area under the curve was 0.829 (Figure 1).

**Table 2.** Identification of Optimal Cut-off Values for Different Predictive Factors Based on the ROC Curve.

| Group | HCC vs HV |  |  | HBV vs HV |  |  | HCC vs HBV |  |  |
| --- | --- | --- | --- | --- | --- | --- | --- | --- | --- |
| Parameter | Cut off | AUC | Youden | Cut off | AUC | Youden | Cut off | AUC | Youden |
| PLR | 174.46 | 0.562 | 0.217 | 75.82 | 0.60 | 0.187 | 129.73 | 0.624 | 0.239 |
| PAR | 327.27 | 0.510 | 0.238 | 150.98 | 0.68 | 0.345 | 229.10 | 0.624 | 0.273 |
| BAR | 0.97 | 0.757 | 0.476 | 0.97 | 0.56 | 0.145 | 0.98 | 0.683 | 0.334 |
| LAR | 3.08 | 0.455 | 0.130 | 1.22 | 0.61 | 0.214 | 1.71 | 0.543 | 0.115 |
| LMR | 3.05 | 0.904 | 0.685 | 3.40 | 0.67 | 0.284 | 2.20 | 0.786 | 0.441 |
| MAR | 0.62 | 0.829 | 0.574 | 0.57 | 0.54 | 0.160 | 0.53 | 0.762 | 0.421 |

Notes: The truncation value, AUC and Youden index of each valuable variable were calculated using the receiver operating characteristic curve.
Abbreviations: BAR, ApoB to ApoA1; HCC, Hepatocellular Carcinoma; HV, Healthy Volunteers; HBV, hepatitis B patients; LAR, lymphocyte to ApoA1; LMR, lymphocyte to monocyte; MAR, monocyte to ApoA1; PLR, platelet to lymphocyte; PAR, platelet to ApoA1.

**Figure 1.**
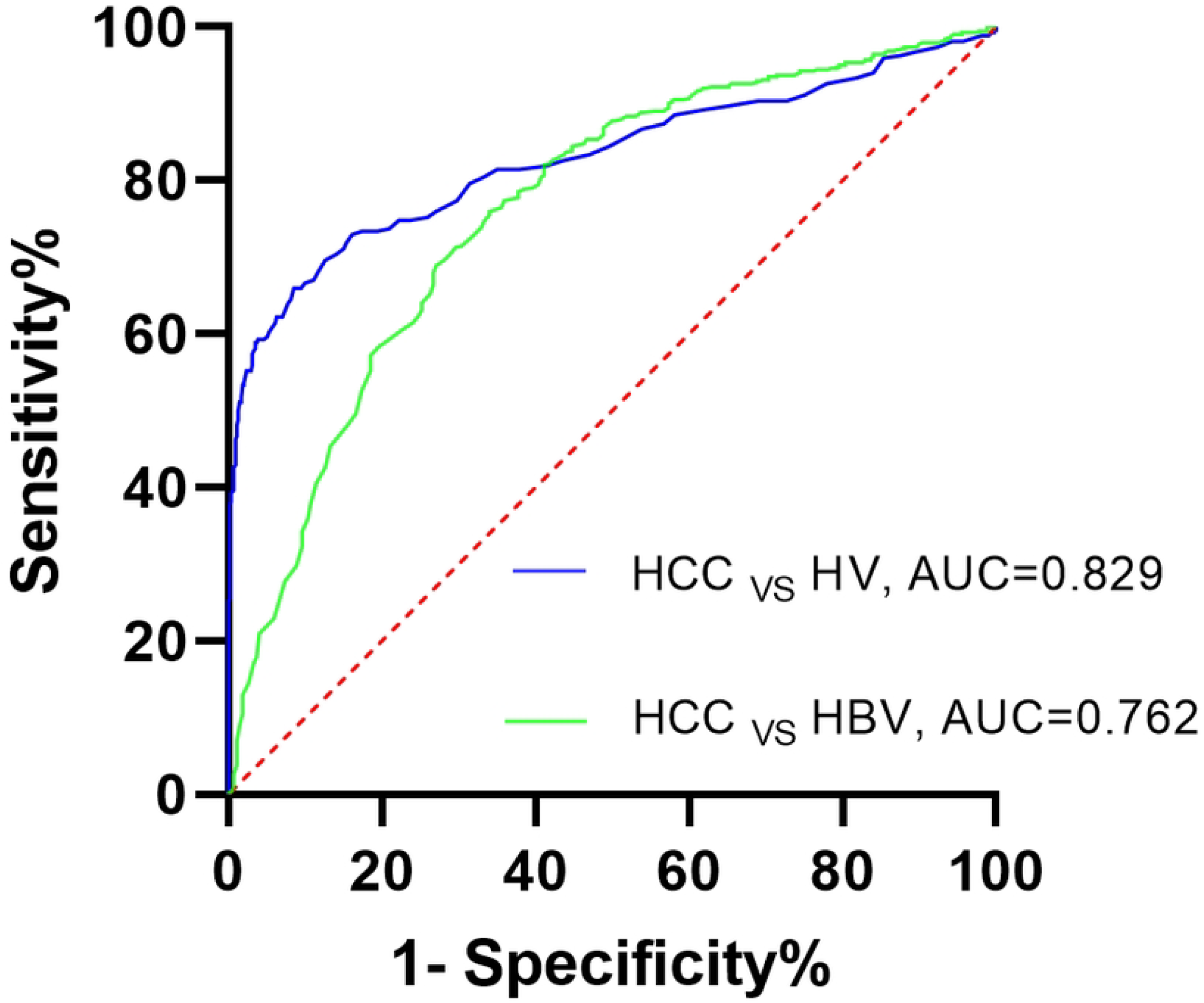
ROC curve analysis of MAR in patients with HCC, Those with hepatitis B patients and Healthy Volunteers.

### Logistic Regression Analysis to Determine Predictive Factors

As shown in Table 3 and 4, univariate and multivariate logistic regression analysis showed that MAR was an indicator to distinguish HCC from hepatitis B disease and healthy people, and also an independent risk factor for HCC. The increase in MAR level demonstrated that the risk of HCC was 6.001 times higher than that of the healthy volunteers (*P* < 0.001).

**Table 3.** The Predictive Factors Identified by Univariate Logistic Regression.

| Group | HCC vs HV |  | HBV vs HV |  | HCC vs HBV |  |
| --- | --- | --- | --- | --- | --- | --- |
| Parameter | OR(95%CI) | *P* | OR(95%CI) | *P* | OR(95%CI) | *P* |
| PLT | 0.986(0.984, 0.988) | 0.000 | 0.986(0.984, 0.988) | 0.000 | 1.000(0.999, 1.002) | 0.654 |
| L | 0.217(0.170, 0.276) | 0.000 | 1.001(0.997, 1.006) | 0.526 | 0.417(0.324, 0.538) | 0.000 |
| M | 12.834(7.237, 22.760) | 0.000 | 0.363(0.201, 0.656) | 0.001 | 18.539(10.203, 33.686) | 0.000 |
| CEA | 1.108(1.042, 1.178) | 0.001 | 1.107(1.040, 1.177) | 0.001 | 1.001(0.998, 1.004) | 0.502 |
| ALB | 0.579(0.547, 0.612) | 0.000 | 0.642(0.609, 0.676) | 0.000 | 0.901(0.880, 0.924) | 0.000 |
| ApoA1 | 0.004(0.002, 0.008) | 0.000 | 0.052(0.032, 0.084) | 0.000 | 0.103(0.065, 0.162) | 0.000 |
| ApoB | 0.371(0.221, 0.622) | 0.000 | 0.302(0.197, 0.462) | 0.000 | 1.147(0.745, 1.767) | 0.532 |
| ALP | 1.025(1.021, 1.029) | 0.000 | 1.010(1.006, 1.014) | 0.000 | 1.015(1.012, 1.018) | 0.000 |
| PLR | 1.004(1.003, 1.006) | 0.000 | 0.999(0.997, 1.000) | 0.097 | 1.004(1.003, 1.006) | 0.000 |
| PAR | 1.002(1.001, 1.003) | 0.000 | 0.998(0.996, 0.999) | 0.000 | 1.003(1.002, 1.004) | 0.000 |
| BAR | 5.539(3.974, 8.085) | 0.000 | 4.092(2.829, 5.920) | 0.000 | 1.346(1.171, 1.547) | 0.000 |
| LAR | 1.051(0.977, 1.130) | 0.180 | 1.052(0.979, 1.132) | 0.168 | 0.999(0.993, 1.004) | 0.625 |
| LMR | 0.386(0.340, 0.440) | 0.000 | 1.001(0.999, 1.002) | 0.520 | 0.499(0.440, 0.567) | 0.000 |
| MAR | 13.229(8.216, 21.303) | 0.000 | 6.573(4.146, 10.421) | 0.000 | 1.981(1.630, 2.409) | 0.000 |

Abbreviations: ALB, albumin; ApoA1, apolipoprotein A1; ApoB, Apolipoprotein B; ALP, alkaline phosphatase; BAR, ApoB to ApoA1; CEA, carcinoembryonic antigen; HCC, Hepatocellular Carcinoma; HV, Healthy Volunteers; HBV, hepatitis B patients; L, lymphocytes; LAR, lymphocyte to ApoA1; LMR, lymphocyte to monocyte; M, Monocytes; MAR, monocyte to ApoA1; PLT, platelet; PLR, platelet to lymphocyte; PAR, platelet to ApoA1.

**Table 4.** The Predictive Factors Identified by Multivariate Logistic Regression.

| Group | HCC vs HV |  | HBV vs HV |  | HCC vs HBV |  |
| --- | --- | --- | --- | --- | --- | --- |
| Parameter | OR(95%CI) | *P* | OR(95%CI) | *P* | OR(95%CI) | *P* |
| PAR | 0.995(0.994, 0.997) | 0.000 | 0.991(0.989, 0.992) | 0.000 | 1.001(1.000, 1.003) | 0.015 |
| BAR | 6.411(3.844, 10.692) | 0.000 | 10.875(6.576, 17.984) | 0.000 | 0.799(0.669, 0.954) | 0.013 |
| LMR | 0.466(0.398, 0.545) | 0.000 | 1.000(0.998, 1.003) | 0.696 | 0.615(0.529, 0.715) | 0.000 |
| MAR | 6.001(2.432, 14.811) | 0.000 | 2.492(1.297, 4.788) | 0.006 | 3.941(1.869, 8.307) | 0.000 |

Abbreviations: BAR, ApoB to ApoA1; HCC, Hepatocellular Carcinoma; HV, Healthy Volunteers; HBV, hepatitis B patients; LMR, lymphocyte to monocyte; MAR, monocyte to ApoA1; PAR, platelet to ApoA1.

### Relationship Between MAR and Other Laboratory Indicators of HCC

As shown in Figure 2, MAR was correlated with L (r = -0.075, *P* < 0.05), CEA (r = 0.171, *P* < 0.001), ALB (r = -0.445, *P* < 0.001), ALP (r = 0.340, *P* < 0.001), PAR (r = 0.463, *P* < 0.001), BAR (r = 0.554, *P* < 0.001), LAR (r = 0.485, *P* < 0.001), LMR (r = -0.653, *P* < 0.001).

**Figure 2.**
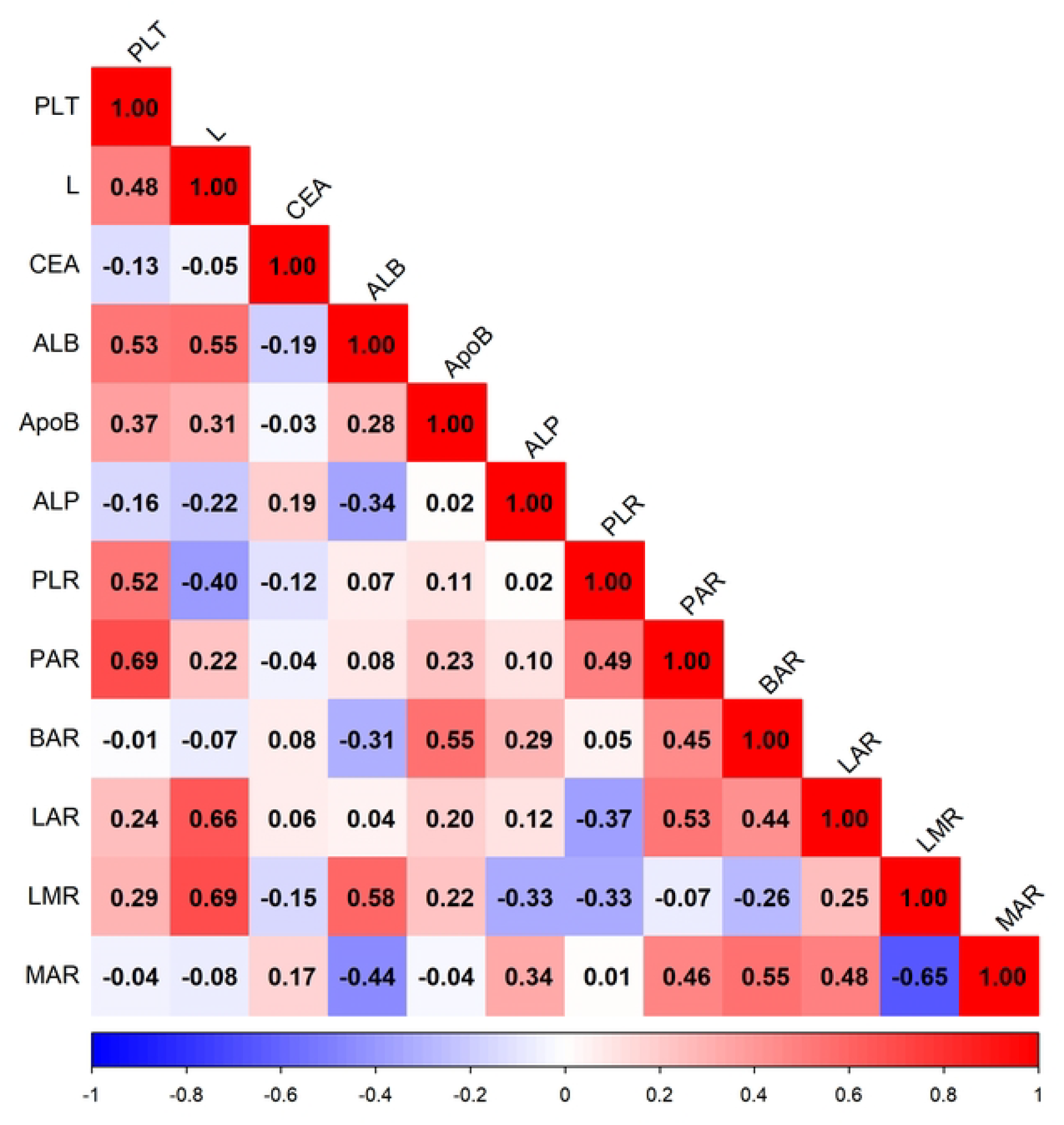
correlation heatmap of Selected Variables.

## Discussion

In this study, the venous blood indicators of HCC, hepatitis B and healthy volunteers were analyzed, and it was found that the level of MAR was the largest in the HCC group, followed by hepatitis B, and the lowest among the healthy volunteers, suggesting that MAR can be used for the auxiliary differential diagnosis of hepatitis B and HCC. It is speculated that the difference of MAR ratio in HCC and hepatitis B diseases is mediated by abnormal blood lipid and chronic inflammation in patients.

ApoA1 as an indispensable component of HDL, inhibits monocyte chemotaxis and recruitment and was shown to be involved in inflammatory reactions, and ApoA1/HDL can be activated and participate in antitumor activities in mature immune systems[17,18]. It was reported that ApoA1 could potently suppress tumor growth and metas□tasis in vivo and improve survival in mouse tumor models[19,20]. Mononuclear cells are the main inflammatory cells in tumor stroma. It has been reported that the increase in activated monocytes [human leukocyte antigen (HLA)-DR^high^CD68^+^ cells] in the liver is related to disease progression[21]. Chemokine (C-C motif) ligand 15 (CCL15) recruits CCR1^+^CD14^+^ monocytes to the edge of HCC tissue. High expression of CCL15 is associated with poor clinical prognosis. CCR1^+^CD14^+^ monocytes suppress antitumor immunity, facilitate tumor metastasis and promote tumor cell proliferation and invasion[22].

Correlation analysis reveals a correlation between MAR and L, CEA, ALB, ALP, PAR, BAR, LAR, LMR. Lymphocytes play a pivotal role in immunosurveillance and immune editing, and lymphocyte infiltration in the tumor microenvironment (TME) contributes to the immunologic anticancer reaction[23,24]. The presence of tumor-infiltrating lymphocytes is associated with improved survival in various cancers, and conversely, low lymphocyte counts and failure to infiltrate the tumor lead to inferior survival[25,26]. LMR is the ratio of lymphocyte to monocyte. Various investigations have shown that a decreased pretreatment LMR has an unfavorable impact on OS in cancer patients among various tumor subgroups[27], including HCC[28,29]. PAR is the ratio of platelet to ApoA1. Platelets facilitate tumor progression by supporting cancer stem cells, induc□ing angiogenesis, sustaining cell proliferation, and evading immune surveillance[30]. In addition platelets might represent a potential therapeutic target, and indeed, sorafenib is a standard of care in HCC treatment directly targeting pathways mediated by VEGF and platelet□derived growth factor (PDGF)[31]. Moreover, previous investigations highlighted the inhibiting effect on tumor growth exerted by antiplatelet therapies (aspirin, warfarin, and cyclo-oxygenase inhibitors), even in HCC[32,33]. All in all, these indicators are all related to inflammation and inflammation-based scores may be convenient, easily obtained, low-cost, and reliable biomarkers with diagnostic and prognostic significance for HCC. Using univariate logistic regression analysis, it was found that MAR is an independent risk factor of HCC, and the increased level of MAR indicated that the risk of HCC is 13.229 times higher than that of healthy volunteers, and multivariate logistic regression analysis found that the risk of HCC is 6.001 times higher than that of healthy volunteers. This ratio is higher than that reported for breast cancer (3.733 times)[16] and Type 2 diabetes mellitus (2.26 times)[15], suggesting that the MAR indicator may have good clinical application prospects.

Wang et al[15] believed that MAR was a risk factor for type-2 diabetes metabolic syndrome. The MAR level of diabetes patients with metabolic syndrome was higher than that of patients without metabolic syndrome. Moreover, Lin et al[16] found that MAR is a new indicator for the differential diagnosis of benign and malignant breast diseases, and also an independent risk factor for breast cancer. High MAR is closely related to late stages of breast cancer, deeper tumor invasion, and distant metastasis. The results of this study showed that high MAR was an independent risk factor for HCC, with a cutoff value of 0.62, high MAR was a risk factor for HCC. In addition, MAR level was the highest in the HCC (0.92), followed by hepatitis B disease (0.38), and the lowest in the healthy volunteer (0.36), MAR may serve as an indicator for predicting the progression from hepatitis B to HCC.

This study has some limitations. This was a retrospective study without follow-up, and it cannot directly reflect the associations MAR and HCC. The significance of the MAR on survival prognosis in patients with HCC needs to be confirmed in a large-scale prospective cohort study. Moreover, the subjects included in this study are local residents, and the optimal cutoff values of MAR may be not applicable to other races.

## Conclusions

The results of this study showed that MAR is a new indicator for the differential diagnosis of HCC and hepatitis B disease, and also an independent risk factor for HCC. To our knowledge, this is the first study to investigate the clinical value of MAR in HCC in China. It is hoped that more research in the future will explore the exact pathogenic mechanism of HCC in this direction, provide reference for the diagnosis and the treatment of HCC, and contribute to the improved quality of life of patients.

## Data Availability

All relevant data are within the manuscript and its Supporting Information files.

## Author contributions

**Conceptualization:** Cheng Su, Fu Wei.

**Data curation:** Cheng Su, Xuefeng Zhu.

**Formal analysis:** Xuefeng Zhu, Fu Wei.

**Funding acquisition:** Ling Li.

**Investigation:** Cheng Su, Xuefeng Zhu, Fu Wei, Ling Li.

**Methodology:** Cheng Su, Fu Wei.

**Project administration:** Zhongyuan Lin, Fu Wei, Ling Li.

**Resources:** Cheng Su, Ling Li.

**Software:** Xuefeng Zhu, Fu Wei.

**Writing – original draft:** Cheng Su, Xuefeng Zhu.

**Writing – review & editing:** Zhongyuan Lin, Fu Wei, Ling Li.

